# Lessons learned from evaluating the NFDI4Health metadata schema for health studies using the Research Data Alliance’s FAIR data maturity model

**DOI:** 10.1101/2025.09.26.25336637

**Authors:** Elisa Kasbohm, Esther Thea Inau, Atinkut Alamirrew Zeleke, Kirubel Biruk Shiferaw, Haitham Abaza, Vera Clemens, Martin Golebiewski, Marisabel Gonzalez-Ocanto, Sophie A. I. Klopfenstein, Carina N. Vorisek, Dagmar Waltemath, Carsten Oliver Schmidt

## Abstract

**Objectives:** The German National Research Data Infrastructure for Personal Health Data (NFDI4Health) has developed a metadata schema (MDS) for harmonizing health study descriptions, along with a platform for content discovery. This work evaluates how well the MDS promotes FAIR data principles (Findability, Accessibility, Interoperability, Reusability) and serves as a proxy for assessing study FAIRness.

**Materials and Methods:** Using the Research Data Alliance’s ‘FAIR data maturity model’, we assessed the scope of evaluable FAIRness indicators for NFDI4Health’s MDS (version 3.3).

**Results:** 29 of 41 FAIRness indicators can be evaluated (Findability: 7/7, Interoperability: 7/12, Accessibility: 7/12, Reusability: 8/10). The remaining indicators relate to the research data’s format and access procedures.

**Discussion:** The MDS provides a viable basis for evaluating study FAIRness. To enable full indicator coverage, additional metadata elements should be incorporated.

**Conclusion:** NFDI4Health’s MDS effectively promotes FAIR-compliant study descriptions, although adjustments could strengthen its utility for assessing research data FAIRness.

**Lay summary:** Making research data easily Findable, Accessible, Interoperable, and Reusable (FAIR) is crucial for accelerating discoveries in health research. The German National Research Data Infrastructure for Personal Health Data (NFDI4Health) is an initiative and network that develops services, tools, and training programs to promote and support the FAIR sharing of health research data in Germany. As part of this effort, NFDI4Health has created a standardized framework, referred to as metadata schema (MDS), to systematically collect and structure information about health studies. The collected information can be accessed through a dedicated platform provided by NFDI4Health. In light of ongoing developments of the platform, it is important to assess whether the MDS content may serve as a proxy for assessing the FAIRness of the included studies. Therefore, we assessed the scope of evaluable FAIRness indicators based on the ‘FAIR data maturity model’ by the Research Data Alliance. Our assessment shows that 29 of 41 FAIRness indicators can currently be evaluated. The remaining indicators relate to the data format and access procedures. However, they could be easily covered by introducing new elements to the MDS and encouraging data holders to provide the required information. Overall, the MDS effectively supports FAIR-compliant descriptions of health studies.

## INTRODUCTION

Metadata repositories play an important role in sustainable medical research. By providing information on the design and content of studies, they foster scientific collaborations and reuse of research data [1]. Various metadata repositories have been established for different fields of medical research. Notable examples for international metadata repositories include ClinicalTrials.gov as register for clinical trials [2], the Maelstrom Catalogue for epidemiological studies [3,4], the euCanSHare platform for cardiovascular research [5,6], and Orphanet for rare diseases [7,8], amongst others. In addition, there are national repositories – in Germany, for example, the German Clinical Trials Register [9] for clinical trials and NFDI4Health’s Health Study Hub for health studies conducted in Germany [10–12].

To maximize the benefits of such metadata repositories, it is helpful to adopt a framework that enables the assessment of the scope and usability of the information they contain. One prominent framework are the FAIR guiding principles [13]. These define a set of criteria to enhance the Findability, Accessibility, Interoperability and Reusability of research data and other scientific outputs. As such, the FAIR principles can guide the development of data sharing infrastructures [13]. The conformance of research data and their representation in a metadata repository with the FAIR principles can be assessed using a list of formal criteria, as for example provided in the ‘FAIR data maturity model’ by the Research Data Alliance (RDA) [14,15]. The ‘FAIR data maturity model’ will be described in more detail below.

The present work is motivated by recent developments within the German National Research Data Infrastructure for Personal Health Data (NFDI4Health), an initiative and network that provides services, tools, and training to enhance the FAIRness of health research conducted in Germany [16,17]. As one of its services, it provides a repository for health studies, including clinical trials, epidemiological studies and public health research, named German Central Health Study Hub (in short: Health Study Hub, HSH) [10,12]. The HSH is based on a metadata schema (MDS) developed by NFDI4Health [18,19], which covers information about health studies such as administrative metadata, research objectives, targeted health conditions or diseases, study design, and information on research data and data sharing. During the development of the MDS, established metadata schemas and standards were considered for compatibility, such as ClinicalTrials.gov [2], the DataCite metadata schema [20], HL7®FHIR® [21], and the MIABIS standard [22–24], amongst others [19,25]. Researchers can enrich the content for their studies in the HSH by uploading and licensing non-sensitive research outputs, such as study protocols, data dictionaries, questionnaires, sample informed consent forms or patient information sheets [11,26]. The HSH allows researchers to publish information about their own studies and to find information about past and ongoing studies using a web interface or JSON-based application programming interfaces (APIs) [10,27]. In addition, NFDI4Health provides software for Local Data Hubs, enabling researchers to store study descriptions and related research outputs locally, with the option of transferring the information to the HSH for public access and reuse [28].

As a means to promote FAIR sharing, NFDI4Health plans to integrate FAIR metrics in their central metadata repository. This requires that the MDS includes all necessary information, which motivates the present assessment of its capabilities.

## OBJECTIVE

The present work evaluates the extent to which NFDI4Health’s MDS (version 3.3), as currently implemented in the HSH, aligns with the FAIR data principles and promotes the publication of FAIR-relevant information for health studies. Ultimately, the evaluation identifies FAIRness indicators that cannot yet be evaluated for studies in the HSH, given the information in the MDS. This is of relevance as it highlights potential limitations in applying FAIR metrics on the studies in the HSH, as well as areas for improvement for harmonized study descriptions in health research.

## MATERIAL AND METHODS

We evaluated the applicability of NFDI4Health’s MDS version 3.3 [18] for FAIRness assessments of its content based on the RDA’s FAIR data maturity model [14,15]. This model comprises 41 indicators that were derived from the FAIR principles [13,14]. The indicators can either be evaluated as ‘fail’ or ‘pass’, or using a rating scale [14]. We chose the former approach, as it is better suited for our feasibility assessment. The assessment was conducted manually through a joint review and evaluation of the indicators involving six authors (EK, ETI, AAZ, KBS, DW, COS). Their consensus was then reviewed by all authors of this work. The results are presented in tabular form.

The indicators of the RDA’s FAIR data maturity model target either ‘data’ or ‘metadata’ [14]. In the context of our assessment, the MDS includes two levels of information: (1) study-level descriptions, referred to as ‘metadata’, which provide details about the study, and (2) metadata describing the properties of the research data generated by the study. The latter enables an assessment of the FAIRness of the underlying ‘data’.

## RESULTS

Tables 1 and 2 report the applicability of the 41 indicators as defined by the RDA, evaluating the current scope of FAIRness indicators included in NFDI4Health’s MDS (version 3.3).

**Table 1.**
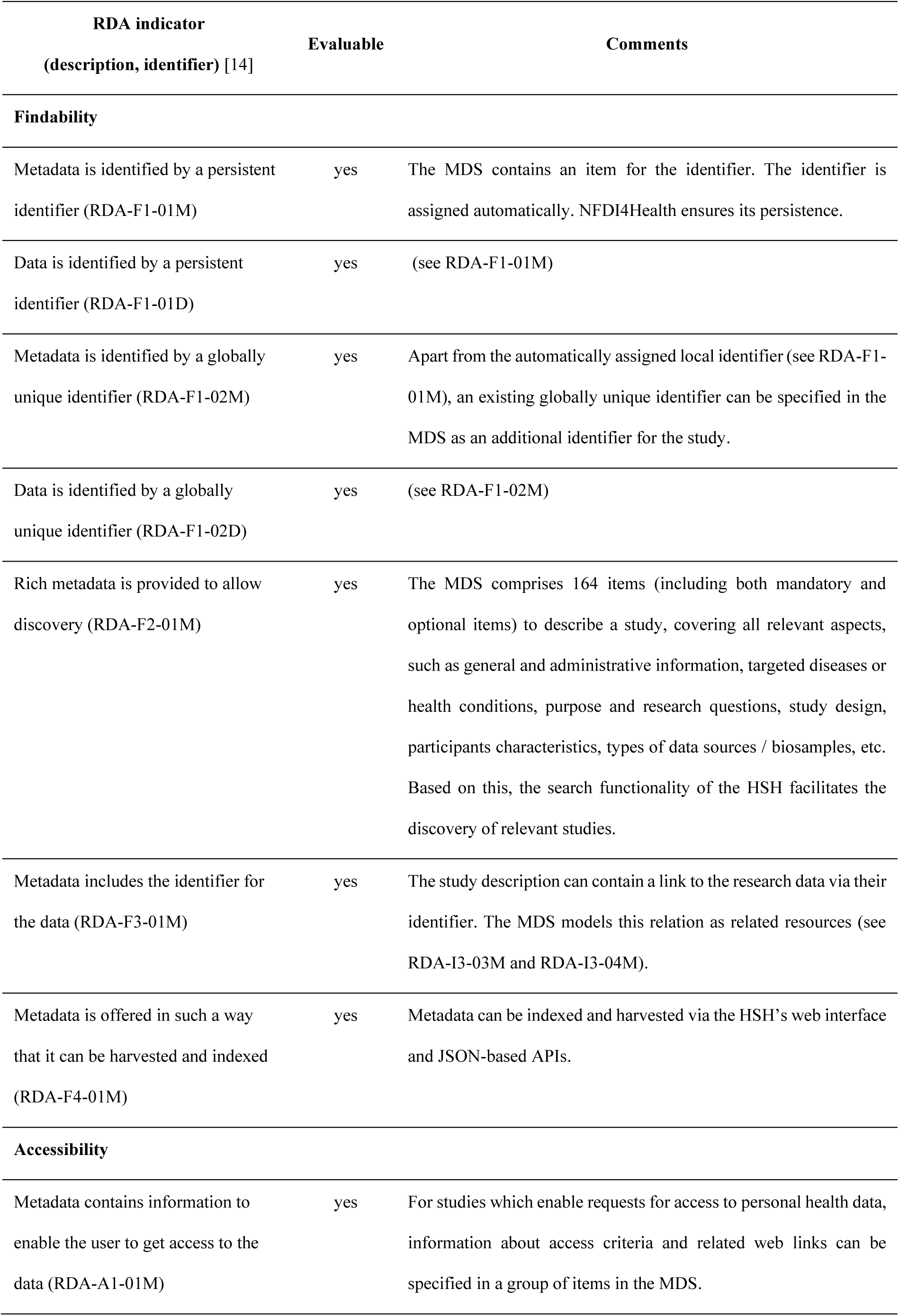
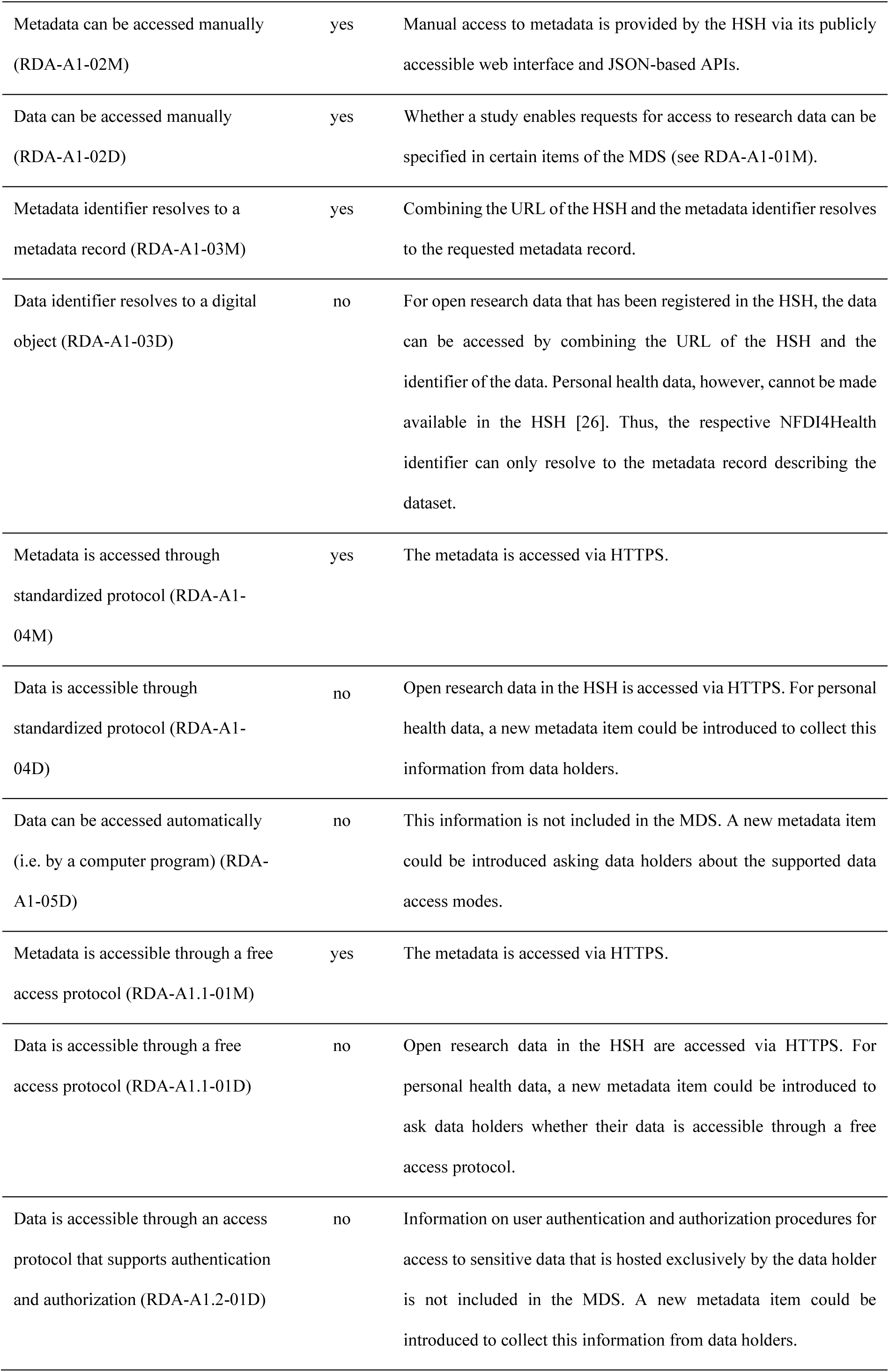
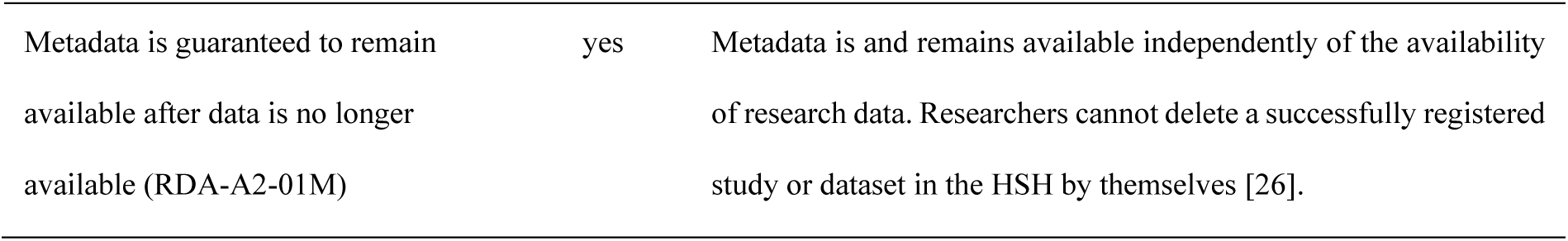
Evaluation of NFDI4Health’s MDS using the RDA’s indicators for Findability and Accessibility.

| **RDA indicator**<br>**(description, identifier) [14]** | **Evaluable** | **Comments** |
| --- | --- | --- |
| **Findability** |  |  |
| Metadata is identified by a persistent identifier (RDA-F1-01M) | yes | The MDS contains an item for the identifier. The identifier is assigned automatically. NFDI4Health ensures its persistence. |
| Data is identified by a persistent identifier (RDA-F1-01D) | yes | (see RDA-F1-01M) |
| Metadata is identified by a globally unique identifier (RDA-F1-02M) | yes | Apart from the automatically assigned local identifier (see RDA-F1-01M), an existing globally unique identifier can be specified in the MDS as an additional identifier for the study. |
| Data is identified by a globally unique identifier (RDA-F1-02D) | yes | (see RDA-F1-02M) |
| Rich metadata is provided to allow discovery (RDA-F2-01M) | yes | The MDS comprises 164 items (including both mandatory and optional items) to describe a study, covering all relevant aspects, such as general and administrative information, targeted diseases or health conditions, purpose and research questions, study design, participants characteristics, types of data sources / biosamples, etc. Based on this, the search functionality of the HSH facilitates the discovery of relevant studies. |
| Metadata includes the identifier for the data (RDA-F3-01M) | yes | The study description can contain a link to the research data via their identifier. The MDS models this relation as related resources (see RDA-I3-03M and RDA-I3-04M). |
| Metadata is offered in such a way that it can be harvested and indexed (RDA-F4-01M) | yes | Metadata can be indexed and harvested via the HSH's web interface and JSON-based APIs. |
| **Accessibility** |  |  |
| Metadata contains information to enable the user to get access to the data (RDA-A1-01M) | yes | For studies which enable requests for access to personal health data, information about access criteria and related web links can be specified in a group of items in the MDS. |
| Metadata can be accessed manually (RDA-A1-02M) | yes | Manual access to metadata is provided by the HSH via its publicly accessible web interface and JSON-based APIs. |
| Data can be accessed manually (RDA-A1-02D) | yes | Whether a study enables requests for access to research data can be specified in certain items of the MDS (see RDA-A1-01M). |
| Metadata identifier resolves to a metadata record (RDA-A1-03M) | yes | Combining the URL of the HSH and the metadata identifier resolves to the requested metadata record. |
| Data identifier resolves to a digital object (RDA-A1-03D) | no | For open research data that has been registered in the HSH, the data can be accessed by combining the URL of the HSH and the identifier of the data. Personal health data, however, cannot be made available in the HSH [26]. Thus, the respective NFDI4Health identifier can only resolve to the metadata record describing the dataset. |
| Metadata is accessed through standardized protocol (RDA-A1-04M) | yes | The metadata is accessed via HTTPS. |
| Data is accessible through standardized protocol (RDA-A1-04D) | no | Open research data in the HSH is accessed via HTTPS. For personal health data, a new metadata item could be introduced to collect this information from data holders. |
| Data can be accessed automatically (i.e. by a computer program) (RDA-A1-05D) | no | This information is not included in the MDS. A new metadata item could be introduced asking data holders about the supported data access modes. |
| Metadata is accessible through a free access protocol (RDA-A1.1-01M) | yes | The metadata is accessed via HTTPS. |
| Data is accessible through a free access protocol (RDA-A1.1-01D) | no | Open research data in the HSH are accessed via HTTPS. For personal health data, a new metadata item could be introduced to ask data holders whether their data is accessible through a free access protocol. |
| Data is accessible through an access protocol that supports authentication and authorization (RDA-A1.2-01D) | no | Information on user authentication and authorization procedures for access to sensitive data that is hosted exclusively by the data holder is not included in the MDS. A new metadata item could be introduced to collect this information from data holders. |
| Metadata is guaranteed to remain available after data is no longer available (RDA-A2-01M) | yes | Metadata is and remains available independently of the availability of research data. Researchers cannot delete a successfully registered study or dataset in the HSH by themselves [26]. |

**Table 2.** Evaluation of NFDI4Health’s MDS using the RDA’s indicators for Interoperability and Reusability.

| **RDA indicator**<br>**(description, identifier) [14]** | **Evaluable** | **Comments** |
| --- | --- | --- |
| **Interoperability** |  |  |
| Metadata uses knowledge representation expressed in standardized format (RDA-I1-01M) | yes | NFDI4Health developed the MDS to standardize study descriptions in health research. The MDS reuses metadata items from related collections and standards, e.g., ClinicalTrials.gov and MIABIS, and controlled vocabularies for specific value sets, e.g., ICD-10 [18,19]. |
| Data uses knowledge representation expressed in standardized format (RDA-I1-01D) | no | This information is not included in the MDS. A new metadata item could be introduced asking data holders whether their data conforms to a standardized format. |
| Metadata uses machine-understandable knowledge representation (RDA-I1-02M) | yes | An API enables users to enter and retrieve the structured study metadata in JSON format from the HSH [27] (see RDA-I1-01M). |
| Data uses machine-understandable knowledge representation (RDA-I1-02D) | no | This information is not included in the MDS. A new metadata item could be introduced asking data holders whether their data use a machine-understandable knowledge representation. |
| Metadata uses FAIR-compliant vocabularies (RDA-I2-01M) | yes | The MDS uses FAIR-compliant vocabularies, such as DataCite and ICD-10. |
| Data uses FAIR-compliant vocabularies (RDA-I2-01D) | no | This information is not included in the MDS. A new metadata item could be introduced asking data holders whether their data uses FAIR-compliant vocabularies. |
| Metadata includes references to other metadata (RDA-I3-01M) | yes | Related metadata objects can be specified in a dedicated group of items in the MDS (see RDA-I3-03M). |
| Data includes references to other data (RDA-I3-01D) | no | This information is not included in the MDS. This aspect might be less relevant, but if it is considered useful, a new metadata item could be introduced to ask data holders whether their data includes references to other data. |
| Metadata includes references to other data (RDA-I3-02M) | yes | Related data can be specified in a dedicated group of items in the MDS (see RDA-I3-04M). |
| Data includes qualified references to other data (RDA-I3-02D) | no | This information is not included in the MDS. This aspect might be less relevant, but if it is considered useful, a new metadata item could be introduced to ask data holders whether their data includes qualified references to other data. |
| Metadata includes qualified references to other metadata (RDA-I3-03M) | yes | The relationship to another metadata object is described in the MDS according to the item 'relationType' by DataCite [16], including its type and identifier, by which the object is linked. |
| Metadata includes qualified references to other data (RDA-I3-04M) | yes | The relationship to other data can be described in the MDS as defined by the item 'relationType' by DataCite [16], including their type and identifier, by which the data is linked. |
| **Reusability** |  |  |
| Plurality of accurate and relevant attributes are provided to allow reuse (RDA-R1-01M) | yes | The current version of the MDS includes many relevant items to describe a study and its research data in detail. Additional modules enable a more precise representation of specific study types, for example in nutritional epidemiology, with further modules being under development, for example for radiomics. |
| Metadata includes information about the license under which the data can be reused (RDA-R1.1-01M) | yes | Datasets can be assigned a license in the MDS. |
| Metadata refers to a standard reuse license (RDA-R1.1-02M) | yes | Standard licenses are recommended, e.g. CC BY 4.0 for study documents [26]. However, other licenses can be specified as well, which may be necessary if institutional licenses apply, e.g. for research data. |
| Metadata refers to a machine-understandable reuse license (RDA-R1.1-03M) | yes | Machine-understandable standard licenses are recommended, e.g. CC BY 4.0 for study documents [26]. |
| Metadata includes provenance information according to community-specific standards (RDA-R1.2-01M) | yes | The 'provenance' entity of the MDS was derived from ClinicalTrials.gov. Domain-overarching provenance information models for life science data, such as the emerging ISO 23494 series [29–31], are under consideration for a future MDS version. |
| Metadata includes provenance information according to a cross-community language (RDA-R1.2-02M) | yes | The MDS contains a 'provenance' entity, which was derived from ClinicalTrials.gov. Cross-community languages for the provenance information, e.g., PROV-O, are under consideration for a future MDS version. |
| Metadata complies with a community standard (RDA-R1.3-01M) | yes | NFDI4Health's MDS serves as a health domain-overarching community standard and was developed in reference to related metadata repositories and standards [19]. |
| Data complies with a community standard (RDA-R1.3-01D) | no | This information is not included in the MDS. A new metadata item could be introduced to ask data holders whether their data complies with a community standard. |
| Metadata is expressed in compliance with a machine-understandable community standard (RDA-R1.3-02M) | yes | Study metadata can be added to and retrieved from the HSH in JSON format, and the MDS is modeled in FHIR [27,32] (see also RDA-R1.3-01M). |
| Data is expressed in compliance with a machine-understandable community standard (RDA-R1.3-02D) | no | This information is not included in the MDS. A new metadata item could be introduced to ask data holders whether their data complies with a machine-understandable community standard. |

In total, 29 out of 41 indicators can be evaluated for study descriptions structured according to NFDI4Health’s MDS, whereas twelve indicators are currently not evaluable. These twelve indicators are all directly related to the research data, specifically regarding details on the data format and data access procedures.

All indicators for Findability can be evaluated (7/7). For Accessibility, seven indicators are evaluable (7/12), whereas five indicators for data access are not covered (RDA-A1-03D, RDA-A1-04D, RDA-A1-05D, RDA-A1.1-01D, RDA-A1.2-01D). Interoperability assessments are currently restricted to seven indicators (7/12), as all five indicators for Interoperability of data are not assessable (RDA-I1-01D, RDA-I1-02D, RDA-I2-01D, RDA-I3-01D, RDA-I3-02D). Eight indicators for Reusability can be assessed (8/10). Only two Reusability indicators are not covered, which target the format of the research data (RDA-R1.3-01D, RDA-R1.3-02D).

## DISCUSSION

Based on the Research Data Alliance’s FAIR data maturity model, we assessed the scope of evaluable FAIRness indicators for studies described with NFDI4Health’s MDS as implemented in the HSH, in order to evaluate whether the MDS captures the information required to calculate FAIR metrics. The development of the MDS is already at an advanced stage and was conducted with the FAIR principles in mind. As a result, many indicators can already be evaluated, either through the way the MDS has been implemented in the HSH or through the available attributes used to describe the studies. Selected indicators for each of the four FAIR principles will be discussed in more detail below.

### Findability

All Findability indicators can be evaluated based on the information in the MDS. The HSH assigns identifiers for study metadata and open research data, ensuring their persistence (RDA-F1-01M, RDA-F1-01D) and uniqueness within NFDI4Health. It is also possible to specify alternative identifiers, which enables to include existing study identifiers from other repositories. Furthermore, the captured metadata includes essential information about a study that can be indexed and harvested from the HSH, for example using its search functionality, enabling researchers to discover relevant studies in their field (RDA-F2-01M, RDA-F4-01M). For instance, the metadata items for chronic diseases were developed based on a review of publicly available information from relevant studies [33].

### Accessibility

For study metadata all Accessibility indicators can be evaluated. The HSH plays a key role here as it provides free and consistent access to study descriptions (RDA-A1-02M, RDA-A1-03M, RDA-A1-04M, RDA-A1.1-01M). Further, NFDI4Health policy mandates that published study descriptions remain publicly available (RDA-A2-01M). Studies that provide access to research data can specify this information in dedicated metadata items (RDA-A1-01M). However, five out of six indicators for accessible research data cannot be evaluated at present. The missing information on details about data access procedures could be collected by asking data holders to report it through dedicated metadata items.

### Interoperability

The MDS currently supports the evaluation of Interoperability of metadata, but not of research data. Specifically, the Interoperability of study descriptions is supported through (qualified) references to other metadata and data objects (RDA-I3-01M, RDA-I3-02M, RDA-I3-03M, RDA-I3-04M) based on the DataCite schema [20]. For instance, study descriptions in the HSH can include references to study documents, datasets, publications, or follow-up studies, making it easier for researchers to discover and reuse related resources. The development of the MDS also included mappings to established metadata schemas in medical research to facilitate interoperability [19]. However, the MDS could be further improved by extending the current use of standardized formats, such as controlled vocabularies [14] (RDA-I1-01M) and FAIR-compliant vocabularies (RDA-I2-01M). This has been addressed by mapping value sets to international terminologies like SNOMED CT, LOINC, and the NCI thesaurus [25], but limitations arise due to the incorporation of metadata from other repositories [34]. Interoperability of research data cannot be evaluated based on the information in the MDS. To address this limitation, new metadata items could be introduced providing information on data formats and vocabularies in the dataset.

### Reusability

The MDS meets several key requirements to assess Reusability, including the provision of information about reuse licenses (RDA-R1.1-01M, RDA-R1.1-02M, RDA-R1.1-03M) and provenance (RDA-R1.2-01M). Only the assessment of Reusability for research data is limited at present. This could be addressed by adding new metadata items to explicitly state whether the research data’s format conforms to a (machine-understandable) community standard (RDA-R1.3-01D, RDA-R1.3-02D).

The reusability of study metadata is essential for avoiding duplicating efforts, particularly in light of the growing number of initiatives for the reuse of data from medical research and routine clinical care [35], such as the European Health Data Space [36], as well as from other fields of research [37]. The continued development of the MDS will likely involve adaptations to accommodate new schemas and standards (RDA-R1.3-01M), since NFDI4Health aims to connect communities from different areas of medical research in Germany.

### Lessons learned

Prospectively designing FAIR metadata schemas is essential to promote and support sustainable research. Various tools and frameworks have been developed for this purpose and can be employed during the design of metadata schemas (e.g., [14,38,39]). For further FAIRification recommendations, see [40–46].

The results of the FAIR assessment of NFDI4Health’s MDS reflect that its initial development focused on FAIR study metadata. Based on this conceptual framework, the HSH contributes to the FAIR sharing of health studies in Germany by providing a platform for researchers to share harmonized study descriptions. This helps to inform researchers and the interested public about research objectives, methods, results, and potential data access. However, more insight into the FAIRness of the research data could be provided by adapting and extending the MDS accordingly. Our findings may support other initiatives in designing metadata frameworks that enable systematic FAIR assessments of study-related data sources.

## CONCLUSION

The present assessment shows that NFDI4Health’s MDS, as implemented in the HSH, supports the evaluation of 29 of the RDA’s FAIRness indicators and therefore enables FAIRness assessments of studies contained in this repository. However, this can be improved by addressing information gaps on the FAIRness of the research data. Ultimately, the inclusion of FAIR metrics is expected to further promote FAIR sharing in NFDI4Health’s network.

## Data Availability

All data produced in the present work are contained in the manuscript.

## AUTHOR CONTRIBUTIONS

EK: Formal analysis, Writing – original draft, Writing – review & editing, ETI: Formal analysis, Methodology, Writing – review & editing, AAZ: Formal analysis, Methodology, Writing – review & editing, KBS: Formal analysis, Methodology, Writing – review & editing, HA: Formal analysis, Writing – review & editing, VC: Formal analysis, Writing – review & editing, MG: Writing – review & editing, MGO: Formal analysis, Writing – review & editing, SAIK: Formal analysis, Writing – review & editing, CNV: Formal analysis, Writing – review & editing, DW: Methodology, Supervision, Writing – review & editing, COS: Conceptualization, Formal analysis, Methodology, Supervision, Writing – original draft, Writing – review & editing

## REFERENCES

1 David R, Rybina A, Burel J, et al. “Be sustainable”: EOSC-Life recommendations for implementation of FAIR principles in life science data handling. The EMBO Journal. 2023;42:e115008. doi: 10.15252/embj.2023115008

2 U.S. Department of Health and Human Services, National Institutes of Health, National Library of Medicine, et al. ClinicalTrials.gov. Protocol Registration Data Element Definitions for Interventional and Observational Studies. 2024. https://clinicaltrials.gov/policy/protocol-definitions (accessed 10 May 2024)

3 Bergeron J, Doiron D, Marcon Y, et al. Fostering population-based cohort data discovery: The Maelstrom Research cataloguing toolkit. PLOS ONE. 2018;13:e0200926. doi: 10.1371/journal.pone.0200926

4 Maelstrom Research. http://www.maelstrom-research.org/ (accessed 30 May 2024)

5 Bienefeld E. euCanSHare: EU-Canada joint data platform to facilitate multi-study cardiovascular research. Published Online First: 11 December 2018. doi: 10.5281/zenodo.2593568

6 euCanSHare. https://mica.eucanshare.bsc.es/ (accessed 30 May 2024)

7 Rath A, Olry A, Dhombres F, et al. Representation of rare diseases in health information systems: The orphanet approach to serve a wide range of end users. Human Mutation. 2012;33:803–8. doi: 10.1002/humu.22078

8 Orphanet: Research and trials. https://www.orpha.net/en/research-trials/research-projects (accessed 30 May 2024)

9 Federal Institute for Drugs and Medical Devices. German Clinical Trials Register. https://drks.de/search/en (accessed 24 September 2025)

10 Darms J, Clemens V, Gonzalez-Ocanto M, et al. The German Central Health Study Hub – A Service to Find and Publish Clinical, Public Health and Epidemiolocal Studies and Associated Documents. German Medical Data Sciences 2024. 2024;129–37. doi: 10.3233/SHTI240847

11 Darms J, Henke J, Hu X, et al. Improving the FAIRness of Health Studies in Germany: The German Central Health Study Hub COVID-19. Applying the FAIR Principles to Accelerate Health Research in Europe in the Post COVID-19 Era. 2021;78–82. doi: 10.3233/SHTI210818

12 NFDI4Health. Health Study Hub. https://health-study-hub.de/ (accessed 24 September 2025)

13 Wilkinson MD, Dumontier M, Aalbersberg IJ, et al. The FAIR Guiding Principles for scientific data management and stewardship. Sci Data. 2016;3:160018. doi: 10.1038/sdata.2016.18

14 FAIR Data Maturity Model Working Group. FAIR Data Maturity Model. Specification and Guidelines. Published Online First: 25 June 2020. doi: 10.15497/rda00050

15 Bahim C, Casorrán-Amilburu C, Dekkers M, et al. The FAIR Data Maturity Model: An Approach to Harmonise FAIR Assessments. Data Science Journal. 2020;19:41. doi: 10.5334/dsj-2020-041

16 Fluck J, Lindstädt B, Ahrens W, et al. NFDI4Health – Nationale Forschungsdateninfrastruktur für personenbezogene Gesundheitsdaten. Bausteine Forschungsdatenmanagement. 2021;2:72–85. doi: 10.17192/bfdm.2021.2.8331

17 Pigeot I, Ahrens W, Darms J, et al. Making Epidemiological and Clinical Studies FAIR Using the Example of COVID-19. Datenbank Spektrum. 2024;24:117–28. doi: 10.1007/s13222-024-00477-2

18 Abaza H, Shutsko A, Golebiewski M, et al. The NFDI4Health Metadata Schema (V3_3). Published Online First: 2023. doi: 10.4126/FRL01-006472531

19 Abaza H, Shutsko A, Klopfenstein SAI, et al. Toward a Domain-Overarching Metadata Schema for Making Health Research Studies FAIR (Findable, Accessible, Interoperable, and Reusable): Development of the NFDI4Health Metadata Schema. JMIR Medical Informatics. 2025;13:e63906. doi: 10.2196/63906

20 DataCite Metadata Working Group. DataCite Metadata Schema Documentation for the Publication and Citation of Research Data and Other Research Outputs v4.4. Published Online First: 2021. doi: 10.14454/3W3Z-SA82

21 HL7 FHIR v4.0.1 Website. http://hl7.org/fhir/ (accessed 30 March 2022)

22 Merino-Martinez R, Norlin L, van Enckevort D, et al. Toward Global Biobank Integration by Implementation of the Minimum Information About BIobank Data Sharing (MIABIS 2.0 Core). Biopreservation and Biobanking. 2016;14:298–306. doi: 10.1089/bio.2015.0070

23 Eklund N, Engels C, Neumann M, et al. Update of the Minimum Information About BIobank Data Sharing (MIABIS) Core Terminology to the 3rd Version. Biopreservation and Biobanking. 2024;22:346–62. doi: 10.1089/bio.2023.0074

24 Biobanking and Biomolecular Resources - European Research Infrastructure (BBMRI-ERIC). BBMRI-ERIC/miabis GitHub repository. 2024. https://github.com/BBMRI-ERIC/miabis (accessed 10 May 2024)

25 Vorisek CN, Klopfenstein SAI, Löbe M, et al. Towards an Interoperability Landscape for a National Research Data Infrastructure for Personal Health Data. Sci Data. 2024;11:772. doi: 10.1038/s41597-024-03615-3

26 Lindstädt B, Shutsko A, on behalf of the NFDI4Health Consortium and the NFDI4Health Task Force COVID-19. Publication Policy of the National Research Data Infrastructure for Personal Health Data (NFDI4Health) and the NFDI4Health Task Force COVID-19. Published Online First: 2022. doi: 10.4126/FRL01-006431467

27 NFDI4Health. German Central Health Study Hub - Application Programming Interface (API) documentation. https://csh.nfdi4health.de/doc/api (accessed 15 February 2024)

28 Hu X, Abaza H, Hänsel R, et al. NFDI4Health Local Data Hubs Implementing a Tailored Metadata Schema for Health Data. German Medical Data Sciences 2024. 2024;115–22. doi: 10.3233/SHTI240845

29 International Organization for Standardization. Biotechnology — Provenance information model for biological material and data — Part 1: Design concepts and general requirements (ISO/DIS 23494-1). https://www.iso.org/standard/89266.html (accessed 25 September 2025)

30 International Organization for Standardization. Biotechnology — Provenance information model for biological material and data — Part 2: Common Provenance Model (ISO/DIS 23494-2). https://www.iso.org/standard/87714.html (accessed 25 September 2025)

31 International Organization for Standardization. Biotechnology — Provenance information model for biological material and data — Part 3: Provenance of Biological Material (ISO/DIS 23494-3). https://www.iso.org/standard/89236.html (accessed 25 September 2025)

32 Hölter TA, Klopfenstein SAI, Sass J, et al. Implementation Guide of the NFDI4Health Metadata Schema (MDS) Version 3.3. https://simplifier.net/guide/nfdi4health---metadata-schema---implementationguide?version=current (accessed 15 February 2024)

33 Schwedhelm C, Nimptsch K, Ahrens W, et al. Chronic disease outcome metadata from German observational studies – public availability and FAIR principles. Sci Data. 2023;10:868. doi: 10.1038/s41597-023-02726-7

34 Miron L, Gonçalves RS, Musen MA. Obstacles to the reuse of study metadata in ClinicalTrials.gov. Sci Data. 2020;7:443. doi: 10.1038/s41597-020-00780-z

35 Waltemath D, Beyan O, Crameri K, et al. FAIRe Gesundheitsdaten im nationalen und internationalen Datenraum. Bundesgesundheitsbl. 2024;67:710–20. doi: 10.1007/s00103-024-03884-8

36 European Commission. Communication from the Commission - A European Health Data Space: harnessing the power of health data for people, patients and innovation. 2022. https://health.ec.europa.eu/publications/communication-commission-european-health-data-space-harnessing-power-health-data-people-patients-and_en (accessed 15 December 2024)

37 Hartl N, Wössner E, Sure-Vetter Y. Nationale Forschungsdateninfrastruktur (NFDI). Informatik Spektrum. 2021;44:370–3. doi: 10.1007/s00287-021-01392-6

38 Bahim C, Dekkers M, Wyns B. Results of an Analysis of Existing FAIR Assessment Tools. Published Online First: 23 May 2019. doi: 10.15497/rda00035

39 Devaraju A, Huber R, Mokrane M, et al. FAIRsFAIR Data Object Assessment Metrics. Published Online First: 14 April 2022. doi: 10.5281/zenodo.6461229

40 Welter D, Juty N, Rocca-Serra P, et al. FAIR in action - a flexible framework to guide FAIRification. Sci Data. 2023;10:291. doi: 10.1038/s41597-023-02167-2

41 Rocca-Serra P, Gu W, Ioannidis V, et al. The FAIR Cookbook - the essential resource for and by FAIR doers. Sci Data. 2023;10:292. doi: 10.1038/s41597-023-02166-3

42 Cox SJD, Gonzalez-Beltran AN, Magagna B, et al. Ten simple rules for making a vocabulary FAIR. PLOS Computational Biology. 2021;17:e1009041. doi: 10.1371/journal.pcbi.1009041

43 Sinaci AA, Núñez-Benjumea FJ, Gencturk M, et al. From Raw Data to FAIR Data: The FAIRification Workflow for Health Research. Methods Inf Med. 2020;59:e21–32. doi: 10.1055/s-0040-1713684

44 Swertz M, van Enckevort E, Oliveira JL, et al. Towards an Interoperable Ecosystem of Research Cohort and Real-world Data Catalogues Enabling Multi-center Studies. Yearb Med Inform. 2022;31:262–72. doi: 10.1055/s-0042-1742522

45 Inau ET, Sack J, Waltemath D, et al. Initiatives, Concepts, and Implementation Practices of the Findable, Accessible, Interoperable, and Reusable Data Principles in Health Data Stewardship: Scoping Review. Journal of Medical Internet Research. 2023;25:e45013. doi: 10.2196/45013

46 Martínez-García A, Alvarez-Romero C, Román-Villarán E, et al. FAIR principles to improve the impact on health research management outcomes. Heliyon. 2023;9:e15733. doi: 10.1016/j.heliyon.2023.e15733

